# Dysregulated transcriptional responses to SARS-CoV-2 in the periphery support novel diagnostic approaches

**DOI:** 10.1101/2020.07.20.20155507

**Authors:** Micah T. McClain, Florica J. Constantine, Ricardo Henao, Yiling Liu, Ephraim L. Tsalik, Thomas W. Burke, Julie M. Steinbrink, Elizabeth Petzold, Bradly P. Nicholson, Robert Rolfe, Bryan D. Kraft, Matthew S. Kelly, Gregory D. Sempowski, Thomas N. Denny, Geoffrey S. Ginsburg, Christopher W. Woods

## Abstract

In order to elucidate novel aspects of the host response to SARS-CoV-2 we performed RNA sequencing on peripheral blood samples across 77 timepoints from 46 subjects with COVID-19 and compared them to subjects with seasonal coronavirus, influenza, bacterial pneumonia, and healthy controls. Early SARS-CoV-2 infection triggers a conserved transcriptomic response in peripheral blood that is heavily interferon-driven but also marked by indicators of early B-cell activation and antibody production. Interferon responses during SARS-CoV-2 infection demonstrate unique patterns of dysregulated expression compared to other infectious and healthy states. Heterogeneous activation of coagulation and fibrinolytic pathways are present in early COVID-19, as are IL1 and JAK/STAT signaling pathways, that persist into late disease. Classifiers based on differentially expressed genes accurately distinguished SARS-CoV-2 infection from other acute illnesses (auROC 0.95). The transcriptome in peripheral blood reveals unique aspects of the immune response in COVID-19 and provides for novel biomarker-based approaches to diagnosis.

## Main

Our understanding of immune mechanisms driving the varied acute, recovery, and post-infectious manifestations of COVID-19 continues to evolve^1^. Recent work has demonstrated altered mRNA profiles during SARS-CoV-2 infection at the site of infection - in respiratory epithelial cells, BAL, or nasal swab samples - highlighting the dysregulated immune responses at local sites^2–6^. However, the manner in which these signals are modulated (or propagated) beyond the respiratory microenvironment plays a significant role in the ability of the host to control these responses. To define the host peripheral blood transcriptional response in subjects with SARS-CoV-2 infection, we performed RNA sequencing on samples from 46 individuals with PCR-positive, symptomatic SARS-CoV-2 infection, 14 of which were sampled at multiple timepoints. Subjects were enrolled when they presented for clinical care, and the time from symptom onset was recorded for each individual sample collected (range 1-35 days, Tables s1-2). Subjects with COVID-19 were divided based on disease severity and time from symptom onset (early ≤10 days, middle 11-21 days, late >21 days). As comparators, we profiled banked blood samples from patients presenting to the Emergency Department with acute respiratory infection (ARI) due to seasonal coronavirus (n=49), influenza (n=17) or bacterial pneumonia (n=23), and matched healthy controls (n=19). Regardless of time from symptom onset, SARS-CoV-2 infection triggered a robust response in circulating leukocytes that varied based on the length of antecedent illness (Fig 1). At early timepoints (≤10 days of symptoms), the host response of most patients was dominated by upregulation of interferon-response signals that are similar to those described for other common viral ARIs^7–10^ (Fig 2). Most interferon-stimulated genes (ISGs) were generally expressed at a higher level than in healthy subjects but were more muted than seen with seasonal coronaviruses (CoV), and much lower than seen in influenza infection. These transcriptional responses were inversely associated with COVID-19 disease duration and viral load, declining over time more slowly than is seen with other common viral infections^11–13^. While many of these ISGs are tightly co-expressed across seasonal CoV and influenza infections, they exhibited bimodal expression in SARS-CoV-2. Some ISGs (e.g., OAS1/2, XAF1) are upregulated similarly to other infectious states while others were dissociated from the common, conserved ISG response and appeared relatively over (LY6E, OASL, IFI27) or underexpressed (SIGLEC1, IFI44L, RSAD2, LAMP3) in SARS-CoV-2 (Fig 2). Selective inhibition of Type I IFNs by SARS-CoV and MERS^14^ has been well-described, and these observed deviations from what are generally effective interferon responses in seasonal viral infections likely contribute to the overall permissive state underlying the prolonged disease course seen with SARS-CoV-2 here and elsewhere.

**Figure 1.**
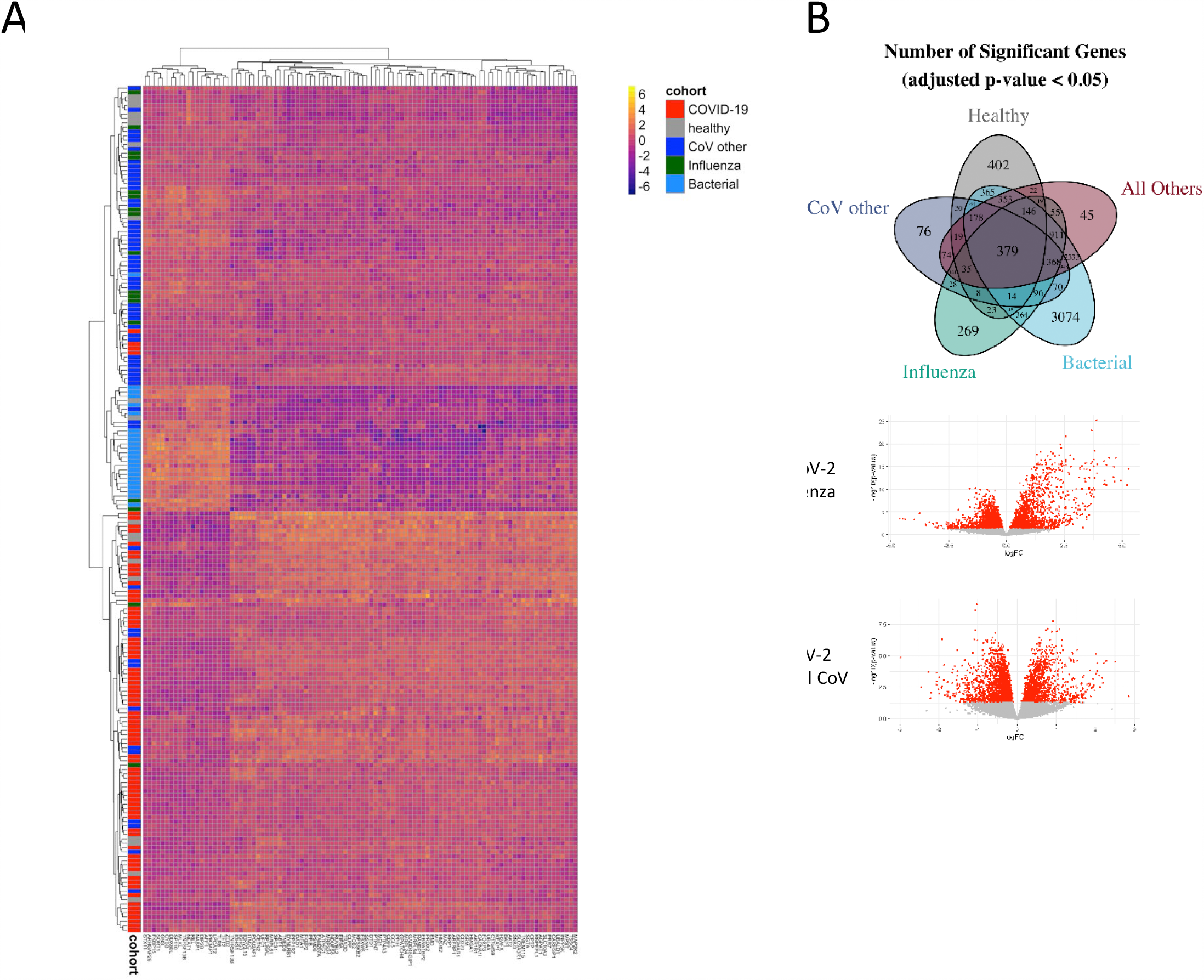
Transcriptomic responses to SARS-CoV-2 in peripheral blood. Heatmap of top differentially expressed genes in COVID-19 versus other infections and healthy controls (A). Venn Diagram of DEGs between COVID-19 and each class, and volcano plot of DEGs in COVID-19 compared to patients with influenza (B, top) and seasonal coronavirus (B, bottom).

**Figure 2.**
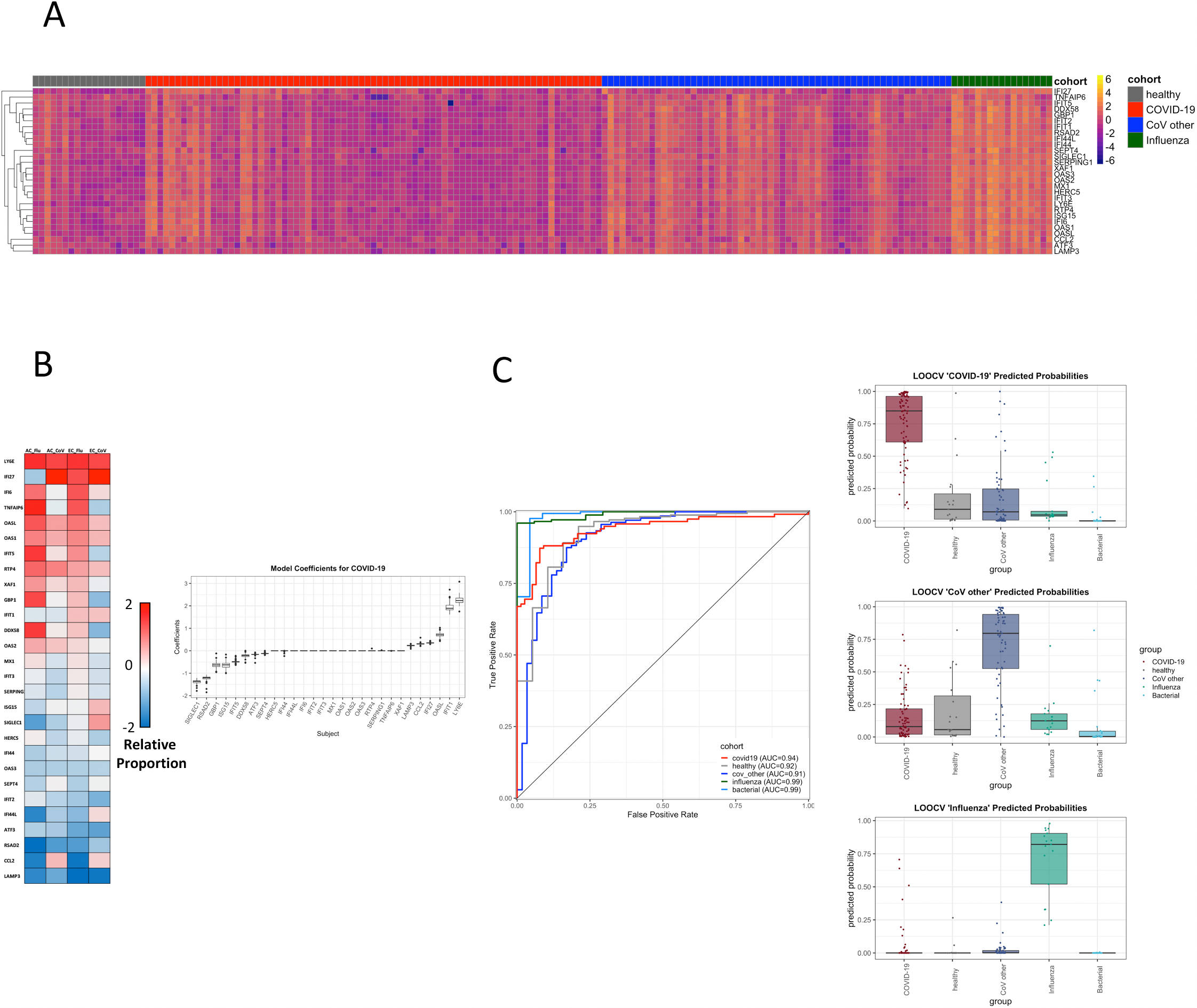
Heatmap of interferon-response based genes from the Interferon-stimulated gene-driven panviral signature across all subjects (A). A number of interferon-stimulated genes are relatively dissociated from one another in Early (EC) or ALL (AC) COVID-19 compared to seasonal coronavirus (CoV) or influenza infections (B, heatmap). A model built on these relative changes utilizes ratios of genes relative to overall ISG signature strength (B, coefficients). A signature built on these interferon-stimulated panviral genes discriminates COVID-19 from others across all time points (C, auROC curve), while the multivariate model built on these genes also simultaneously identifies seasonal CoV and influenza infections (C, right).

Based on these findings, we examined gene expression signatures in peripheral blood consisting primarily of interferon-stimulated genes that have been shown to accurately identify viral ARI across a broad array of seasonal viruses^7,9,10^. One such previously reported ‘panviral’ signature even accurately identifies subjects exposed to respiratory viral pathogens prior to symptoms, often before detectable viral shedding is present^15^. In the current dataset, a regression model built on the 28 genes included in this panviral signature identified the presence or absence of symptomatic SARS-CoV-2 infection with a high degree of accuracy (cross-validated auROC 0.95, Fig 2). Interestingly, with a change in the relative weights and coefficients of the model, measurement of these exact mRNAs can also be utilized to diagnose and differentiate COVID-19, seasonal coronavirus, or influenza infections (Fig 2), consistent with prior work^7,8^.

Diagnostic accuracy was preserved throughout the prolonged course of COVID-19 in these subjects despite heterogeneity of ISG expression over time, in large part due to the unique dissociation of these responses from one another. The potential for a diagnostic approach based on these findings is profound, as they raise the possibility that measurement of expression levels of a single small set of genes could allow both for detection of SARS-CoV-2 infection as well as simultaneously giving information about the presence of other viral (or even bacterial) infections. Also, since signatures built on these genes accurately detect early, pre-symptomatic influenza and seasonal coronavirus infections^15^, it is possible that tests based broadly on this approach may offer similar early detection of COVID-19 in exposed individuals. When combined with emerging nucleic acid detection platforms that offer sample-to-answer times measured in minutes, successful demonstration of pre-symptomatic COVID-19 detection could contribute to real-time outbreak surveillance and quarantine decisions for asymptomatic but potentially contagious hosts that may drive much of the spread of this disease^16^. However, use in such a scenario will need to be properly validated in exposed but asymptomatic subjects with COVID-19.

In addition to altered interferon responses, subjects with early symptomatic COVID-19 exhibited marked upregulation of B-cell activation (CD79 a/b) and a broad diversity of immunoglobulin genes (IGHG1, IGHV2-5, IGHV3.30, IGLV3-19, IGLV3-25 and others) (Fig 3). This distinct transcriptional manifestation of humoral activation occurred as early as one day after symptom onset and was highest during the first 7 days before gradually declining throughout the recovery phase of illness (Fig 3). This signal corresponded to serum IgA expression as early as the first day of clinical disease, specific serum IgG expression by day 8 as seen here and elsewhere^17^, and to an early rise in the proportion of plasmablasts/plasma cells relative to other viral infections (Fig 3). Given these unique, conserved findings we explored utilizing linear regression modeling to develop improved diagnostic signatures including these B-cell pathways that best differentiate COVID-19 from other infections. A second, conserved multivariate transcriptomic signature emerged that differentiated subjects with SARS-CoV-2 infection from all others with a similarly high degree of accuracy (auROC 0.95), regardless of duration of disease or degree of symptoms at the time (Fig 3). This 139-gene signature was capable of simultaneously identifying influenza or seasonal coronavirus infections (auROC 0.98 and 0.97 respectively) and bacterial pneumonia (auROC 0.98). While such findings need to be validated and optimized using sequencing data from additional cohorts of subjects with COVID-19 (once they are available), the discovery of multiple high-performing gene expression-based signatures in peripheral blood based on different aspects of the unique host response to SARS-CoV-2 reinforces the promise of developing stable, reproducible, and flexible point-of-care host response assays to aid in detection and control of COVID-19.

**Figure 3.**
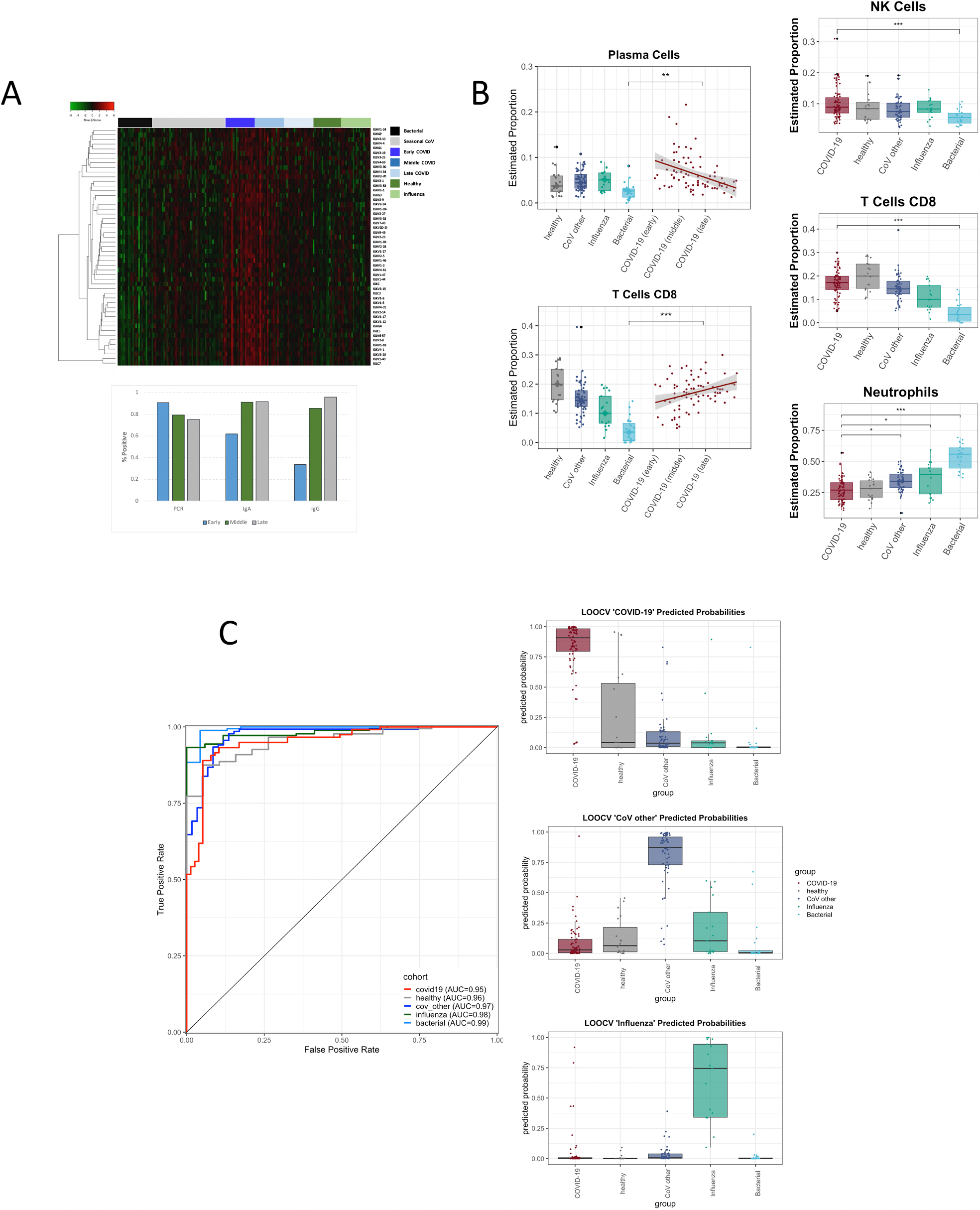
Altered gene expression pathways in COVID-19. Indicators of B-cell activation and Immunoglobulin genes are highly upregulated in SARS-CoV-2 compared to other infections (A, heatmap), which corresponds to early detectable antibody (A, bottom) as well as early elevation of plasmablasts in SARS-CoV-2 patients compare to other infections (B). A gene expression signature comprised of these immunoglobulin and other genes also discriminates SARS-CoV-2 infection at any time from seasonal coronavirus, influenza, and bacterial infections (C).

The data also demonstrated transcriptomic manifestations of additional biological pathways in COVID-19 subjects relevant to clinical disease. SARS-CoV-2 infection is associated with thrombotic events, perhaps due to a hyperinflammatory state^18^, which has led to recommendations for prophylaxis against venous thromboembolism (VTE) in hospitalized patients with COVID-19^19^. Compared to other seasonal CoVs and influenza, we observed marked dysregulation of a genomic signature of VTE^20^ and increased expression of many thrombotic pathway genes in a subset of early COVID-19 cases including prekallikrein (KLKB1), Factor 12 (F12), the plasminogen activator inhibitor (SERPINE1) and others (Fig 4), along with decreased expression of antithrombotic protein S (PROS). This signal was most prominent in early disease but persisted in a subset of individuals for as long as 35 days, and was even more prominent in critically ill subjects. However, further studies of patients with proven thrombotic or microthrombotic disease will be needed to ascertain whether these changes are directly associated with clinical risk of thrombosis. We next examined whether inflammatory pathways targeted by candidate immunomodulatory therapies^21^ demonstrated altered gene expression in these subjects. In a subset of early mild-moderate infections, there was marked dysregulation of IL1 (anakinra), JAK/STAT (baricitinib), IL6 (tocilizumab/sarilumab), and IL10 signaling pathways compared to other infections (Fig 4, Fig s1). In early COVID-19 there is predominantly muted expression of these pathways, where expression levels more closely resemble healthy controls than seasonal coronavirus or influenza, which is consistent with permissive hypoinflammatory responses described elsewhere^5,6^. Severely ill subjects exhibited transcriptional heterogeneity, but showed a trend towards greater IL10 and IFN-response activation along with neutrophil activation, degranulation and translation initiation, but muted IL1 and IL6 signaling. They also demonstrate further elevation in plasmablasts/plasma cells compared to mild disease, but decreased proportions of CD8+ T cells (Fig 4). While this study has low numbers of severely ill subjects, the heterogeneity seen in expression of these pathways across individuals suggests the possibility that pharmacogenomic approaches may prove useful to determine which patients may be most appropriate for a given immunomodulatory agent to mitigate severe disease, and thus should be further explored.

**Figure 4.**
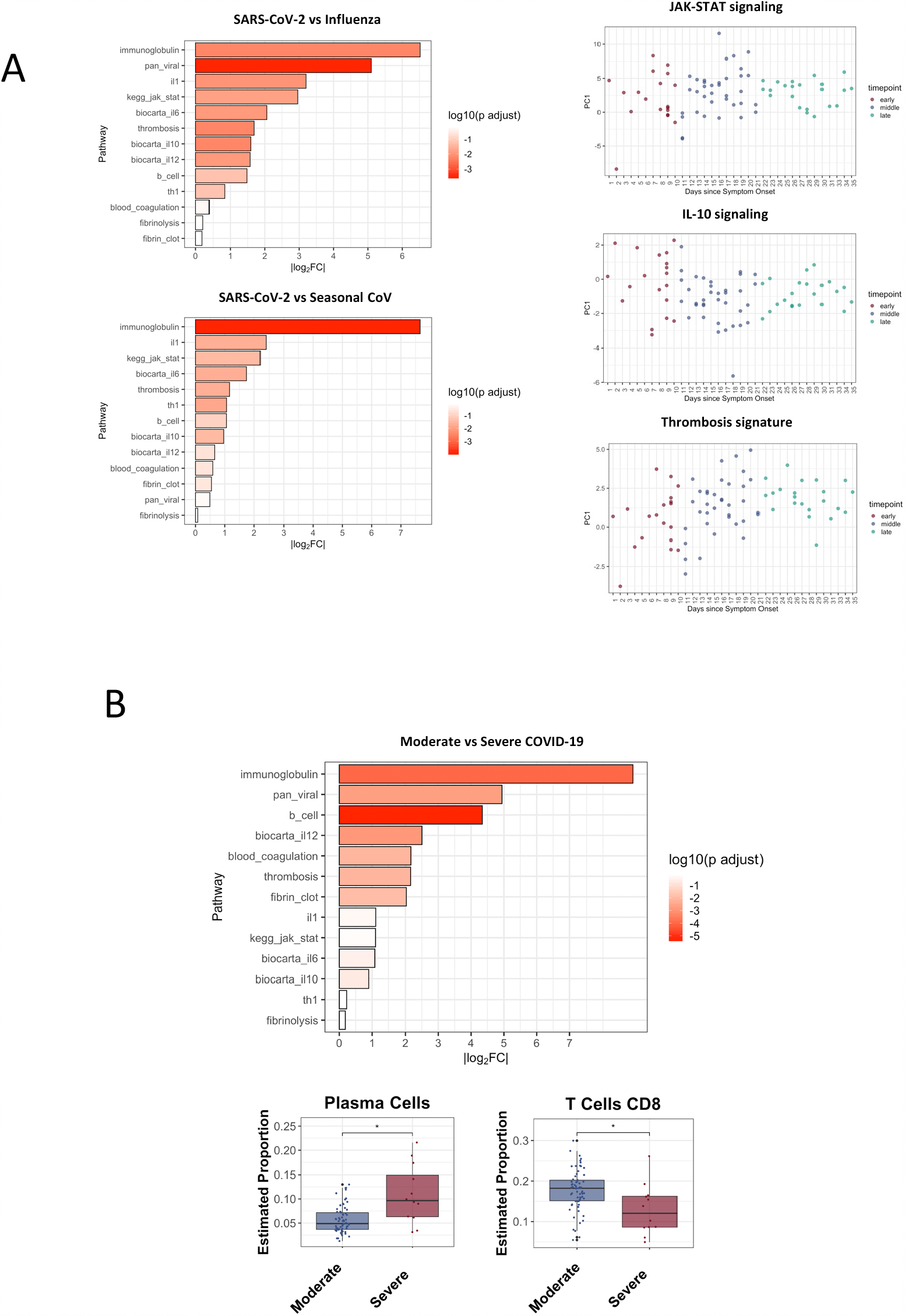
Dysregulated biological pathways in COVID-19. Log2FC and significance of changes in the principle component of relevant biological pathways in subjects with COVID-19 compared to other infections (A). Relative dysregulation of pathways driving IL-10 signaling, JAK-STAT signaling, and a transcriptomic signature of thrombosis over time are also shown (A, right, top to bottom). Similarly, Log2FC change in the principle component of each biological pathway demonstrates differences between moderate and severe COVID-19, with p-values noted for each comparison (C, top) along with relative change in cell type subsets between subjects with moderate and severe COVID-19. (C, bottom).

Together with prior studies^2,5,6,22^, these data show that the transcriptional landscape of the host response to SARS-CoV-2 infection is robust, can elucidate key biological mechanisms of disease, may prove useful for therapeutic drug selection, and contains conserved components which show promise for a new generation of host-based diagnostics to combat this devastating disease.

## Methods

### Institutional Review Board Approvals

The relevant protocols were approved by the IRBs of participating institutions, and were conducted in accordance with the Declaration of Helsinki, applicable regulations and local policies.

### Clinical Cohort Enrollment

Patients with confirmed SARS-CoV-2 infection in either the hospital or outpatient setting were identified through the Duke University Health System (DUHS) or the Durham Veterans Affairs Health System (DVAHS) and enrolled into the Molecular and Epidemiological Study of Suspected Infection (MESSI, Pro00100241). RT-PCR testing for SARS-CoV-2 was performed at either the North Carolina State Laboratory of Public Health or through the clinical laboratory at either DUHS or DVAHS. Subjects with acute respiratory illness of alternative etiologies including seasonal coronavirus, influenza, or bacterial etiologies were prospectively enrolled from a Duke University undergraduate cohort (Predicting Health and Disease, Pro00082317); emergency departments at DUHS, DVAHS, Henry Ford Hospital, or University of North Carolina as part of the CAPSOD (Community-Acquired Pneumonia and Sepsis Outcome Diagnostics, ClinicalTrials.gov NCT00258869), CAPSS (Community-Acquired Pneumonia and Sepsis Study), or RADICAL (Rapid Diagnostics in Categorizing Acute Lung Infection) studies. Written informed consent was obtained from all subjects or legally authorized representatives. All subjects enrolled in CAPSOD, CAPSS, and RADICAL underwent clinical adjudication as previously described^7^. Multiplex viral PCR testing was performed for all subjects using the ResPlex 2•0 viral PCR multiplex assay (Qiagen), xTAG RVP FAST 2 (Luminex), or NxTAG Respiratory Pathogen Panel (Luminex).

### RNA Sequencing

Peripheral blood was collected in PAXgene™ Blood RNA tubes (PreAnalytiX), and total RNA extracted using the PAXgene™ Blood miRNA Kit (Qiagen) employing the manufacturer’s recommended protocol. RNA quantity and quality were assessed using Nanodrop 2000 spectrophotometer (Thermo-Fisher) and Bioanalyzer 2100 with RNA 6000 Nano Chips (Agilent). RNA sequencing libraries were generated using NuGEN Universal mRNA-seq kit with AnyDeplete Globin (NuGEN Technologies, Redwood City, CA) and sequenced on the Illumina NovaSeq 6000 instrument with S4 flow cell and 100bp paired-end reads (performed through the Duke Sequencing and Genomic Technologies Core).

### Statistical Analysis

RNA Sequencing data was normalized using the frozen RMA method ^23^. The sequencing reads were aligned to the human reference genome GRCh38 using STAR^24^ and a count matrix obtained. Genes with counts per million greater than 1 in fewer than 20% of samples were dropped along with three samples with a high proportion of lowly expressed reads. The data was normalized using trimmed mean normalization^25^ and then log2 transformed.

We first performed univariate testing between the COVID-19 cohort and all other cohorts (healthy, Influenza, CoV other, Bacterial), both as COVID-19 against a single other cohort and as COVID-19 against all other groups at once. Additionally, we repeated these analyses with the COVID-19 cohort restricted to the Early COVID-19 samples. Generalized linear models for univariate testing were implemented using the limma package in R^26^. For each comparison, we report multiple hypothesis testing corrected p-values (Benjamini-Hochberg).

Next, we identified differentially expressed pathways between the cohorts of interest. We repeated the above comparisons and perform a similar univariate testing procedure. The normalized expression of the genes in each pathway was summarized as their first Principal Component (PC). These PCs were then used for univariate testing. We computed coordinates of our samples with respect to the first principal component to obtain a dataset of pathway ‘expressions’, exactly analogous to the gene expressions previously tested.

Finally, we trained a statistical model that predicts the cohort label that a subject belongs to. We fit a sparse multinomial logistic regression model to the data^27^ and performed parameter selection and performance estimation via a nested leave-one-out cross validation procedure on the subjects. We used the glmnet package in R^28^ for the basis of our implementation.

Performance was estimated in terms of Area under the Curve (AUC) of the Receiving Operating Characteristic (ROC) for binary comparisons involving COVID-19 vs. other cohorts.

### Analysis of Estimated Cell Type Proportions

The CIBERSORTx method was used to estimate cell-type proportions and perform batch correction for platform differences^29^. A validated signature matrix (LM22) derived from microarray data with 22 human hematopoietic populations was used. Estimated cell types were grouped into primary categories. A linear-mixed model was used to test for differences in etiologies, time-from-symptom onset, and hospital-admission status. The model accounted for the multiple-per-subject measurements. P-values were adjusted for multiple comparisons using the Benjamini–Hochberg method.

## Data Availability

The sequencing datasets generated during and/or analysed during the current study will be made publicly available through the National Center for Biotechnology Information Gene Expression Omnibus.

## Acknowledgements

The authors would like to thank Tiffany Evans, Maria Miggs, Christina Nix, Robert Rolfe, John Bonnewell, Allison Fullenkamp, Versailles Gonzalez, Anna Mazur, T. Scott Alderman, Rosemarie Asrican, and Katherine Frankey, for their tireless work on this study. This work was supported by NIH/NIAID (U01AI066569, UM1AI104681), the U.S. Defense Advanced Projects Agency (DARPA, N66001-09-C-2082 and HR0011-17-2-0069), the Veteran’s Affairs Health System, and Virology Quality Assurance (VQA) # 75N93019C00015. COVID-19 samples were processed under BSL2 with aerosol management enhancement or BSL3 in the Duke Regional Biocontainment Laboratory which received partial support for construction from NIH/NIAID (UC6AI058607).

## Author Statement

MTM, FJC, ELT, GSG, JS and CWW drafted the manuscript, which was critically revised by all remaining authors. MTM, BPN, EP, TB, GSG, RR, and CWW helped conceive and implement the study. All authors helped acquire, analyze, or interpret data. Statistical analyses were performed by FJC, RH, and YL.

